# The Two-Gasometry Indirect Fick Method: An Inconsistent Method for Measuring Cardiac Output in Critical Patients

**DOI:** 10.1101/2025.03.05.25323438

**Authors:** Daniela Cuellar-Mendoza, Javier Mancilla-Galindo, Esmeralda Garza-Santiago, Liliana A. Fernández-Urrutia, Gabriela A. Bautista-Aguilar, Ashuin Kammar-García, Ernesto Deloya-Tomas, Orlando R. Pérez-Nieto

## Abstract

**Background:** The two-gasometry indirect Fick method (iFM) is commonly applied to estimate cardiac output (CO) and cardiac index (CI) in low-resource settings, but validation against accessible methods such as transthoracic echocardiography (TTE) is lacking. We evaluated agreement and clinical interchangeability of CO and CI measurements from TTE (reference test) and two-gasometry iFM (index test) in critically ill patients.

**Methods:** Cross-sectional study conducted in a low-resource critical care unit. CO and CI were measured by intensive care medicine residents using TTE (3 measurements per patient-event) and two-gasometry iFM (1 measurement per patient-event). Agreement was evaluated using mean absolute difference, linear mixed-effects models, and Bland-Altman analysis for repeated measures with 95% limits of agreement (LoA). Precision, bias, and variability were quantified with coefficients of variation, error, and least significant change (LSC), with 95% confidence intervals (95%CI) using bootstrapping.

**Results:** A total of 243 measurements were performed across 81 time points in 52 participants. The two-gasometry iFM showed poor correlation with TTE for both CO (ICC = 0.05) and CI (ICC = –0.04). Mean bias was −2.48 L/min (95% LoA: −8.93 to 3.98) for CO and −1.53 L/min/m² (95% LoA: −5.27 to 2.22) for CI. Mean absolute percentage error was 57.5% (95%CI: 45.5 to 74.6). The LSC was 11.9% for TTE and 80.4% for two-gasometry iFM.

**Conclusions:** The two-gasometry indirect Fick method has poor precision, high variability, and significantly overestimates CO by ∼2.5 L/min and CI by ∼1.5 L/min/m² compared to TTE. Therefore, it is unsuitable for clinical decision-making in critical patients.

**Key points:**

- Question: Is there agreement and clinical interchangeability of CO and CI measurements from transthoracic echocardiography (TTE) and the two-gasometry indirect Fick method (iFM) in critical care patients?
- Finding: The two-gasometry indirect Fick method shows poor agreement with transthoracic echocardiography for measuring cardiac output and cardiac index due to systematic bias, high variability, and lack of precision.
- Meaning: A minimal change of 80.5% in CO with the two-gasometry iFM is needed to be considered a clinically informative change, making this method unsuitable for clinical decision-making in critically ill patients.

## Introduction

Cardiac output (CO) measurement is used for hemodynamic assessment of critically ill patients, as it provides information on cardiac performance and systemic perfusion. The thermodilution technique using a pulmonary artery catheter (PAC)^1^ has been considered the referebce test for CO measurement due to its reliability and direct measurement capability. PACs have been widely used for hemodynamic monitoring in critically ill patients; however, their use is associated with complications including pulmonary artery rupture, arrhythmia, infection, and thrombosis,^2^ driving the development of new technologies. Less invasive or minimally invasive devices have been introduced such as transthoracic echocardiography (TTE), transpulmonary thermodilution,^3^ pulse contour analysis,^4^ estimated continuous cardiac output (esCCO) and bio-reactance-based devices.^5^ Despite their advantages, these are limited by high cost, need for specific consumables, and low availability in resource-limited settings.

TTE has gained popularity due to reduced risks and increased accessibility, in addition to emerging techniques, such as the Fick principle, which offer alternative approaches to CO estimation.^6^ Fick’s principle calculates CO based on the relationship between oxygen consumption (VO□) and the arteriovenous oxygen difference (AVDO□):

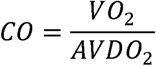

VO□ can be measured directly using a spirometer or Douglas bag,^7^ whereas AVDO□ is determined by arterial and venous blood gas analysis. Nonetheless, this method is rarely implemented as it is resource-intensive and required specialized equipment.

A simplified adaptation, the indirect Fick method (iFM) from two blood sample gas analysis is commonly used in resource-limited settings.^8,9^ Arterial blood is typically drawn from the radial artery, and central venous blood from a catheter placed near the right atrial inlet, where CO is calculated as:

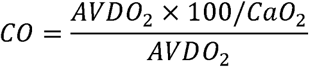

A prospective observational study found no significant correlation between CO measured by thermodilution and formulas based on the Fick method with two blood gas tests.^10^ This raises concerns about the reliability and possible diagnostic errors of the two-gasometry iFM in critically ill patients.

TTE has emerged as a reliable and accessible method due to its strong concordance with PAC. TTE estimates CO indirectly by assessing stroke volume and heart rate.^11^ Multiple studies have shown adequate correlation, acuracy, and precision between CO measured by TTE and thermodilution.^12,13^ Studies in Latin America have shown that TTE and PAC are clinically interchangeable in low-resource settings.^14–16^ Noteworthy, basic echocardiography training permits reliable CO measurements compared to PAC.^17^

Given the resource-intensive and invasiveness of the two-gasometry indirect Fick method, its reliability requires further evaluation. Therefore, we aimed to determine the clinical concordance and interchangeability of CO and cardiac index (CI) measurements obtained by TTE (reference test) and the two-gasometry iFM (index test) in critically ill patients in resource-constrained settings.

## Methods

### Study Design

This cross-sectional study was conducted in the Adult Intensive Care Unit of San Juan del Río General Hospital (Querétaro, Mexico) from June 2021 to December 2023. Inclusion criteria were patients 18 years or older with a pre-indicated central venous catheter. Exclusion criteria included inadequate echocardiographic windows (parasternal long axis, apical 4- and 5-chamber views), no indication for arterial blood gas analysis, positive Allen test, uncontrolled arrhythmia, severe valvular disease, pregnancy, or any condition hindering TTE measurements, as well as cases where CO or CI calculations via TTE or Fick could not be validated analytically.

Out of 67 eligible participants, eleven were excluded due to poor TTE windows and four due to inability to corroborate the CO and CI measurements, yielding 52 participants for analysis. TTE and the two-gasometry iFM were performed at a single time point for most participants, with with ∼25% undergoing repeated assessments to model intra- and inter-subject variance;^18^ three TTE readings were obtained per time point to assess reference test precision.^19^ Repeat measurements occurred 12–48 hours apart, and changes in inotropic or vasopressor use were recorded.

Ventilator parameters recorded at each CO measurement included tidal volume, respiratory rate, PEEP, FiO□, and mode. Lung-protective strategies were applied: 6–10 mL/kg predicted body weight for patients without ARDS, and 4–8 mL/kg for those with ARDS.

All procedures adhered to institutional ethical standards and the Helsinki Declaration (1975). Written informed consent was obtained. The study was approved by the ethics and research committee of the Queretaro Health Services on 14/02/2024 under study title *Método-de-Fick-vs-ecocardiografia-funcional-como-método-de-monitoreo-del-gasto-cardiaco-en-pacientes-de-la-unidad-de-cuidados-intensivos-del-Hospital-General-San-Juan-del-Rio* (registration number: 1616/08/09/2024).

### Cardiac Output Determinations

#### Transthoracic Echocardiography

Two intensive care medicine fellows performed TTE measurements following the technique recommended by the World Alliance of Societies of Echocardiography.^20^ Their training included over 60 hours of continuing medical education in critical care ultrasound, more than 50 supervised echocardiographic studies, a 30-hour course on ultrasound in critical care resuscitation, and one month of hands-on experience in a specialized Cardiology ICU. This exceeds the 30 supervised studies suggested by international guidelines as a reasonable benchmark for competency in image acquisition.^21,22^ The training level of the fellows aligns with the Basic Level classification of the Australian and New Zealand framework.^23^

A total of three TTE measurements were conducted at every time point. The result of each measurement and its average was registered. Cardiac output was measured as the product of stroke volume (SV) and heart rate (HR) as *CO* = *SV* × *HR*.^11^ The stroke volume (SV) was calculated as *SV* = *LVOTa* × *VTI*, where *VTI* is the velocity time integral. The left ventricular outflow tract area (*LVOTa*), also named cross-sectional area (CSA), was calculated as *LVOTa* = π(*d*/2)^2^, where *d* is the left ventricular outflow tract diameter (*LVOTd*). The stroke volume (SV) was calculated as *SV* = *LVOTa* × *VTI*, where VTI is the velocity time integral. The cardiac index (CI) was computed with the formula *CI* = *CO*/*BSA*, where the body surface area is BSA = 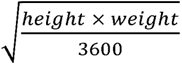.

Ultrasound evaluations were performed with patients in a supine position and the head elevated at 30°. A sector probe (3.5–5 MHz) on the G&E Venuo Go™ ultrasound was used. LVOT diameter was measured in mid-systole from the parasternal long axis view, using inner-to-inner border landmarks or between the insertion points of the right and noncoronary aortic leaflets. LVOT VTI was obtained from the apical five-chamber view using pulsed wave Doppler (PWD), with the sample volume placed ∼1 cm below the aortic valve. An optimal VTI required alignment of the Doppler beam parallel to subaortic flow, or within 30°, with minimal spectral broadening.

#### Two-gasometry Indirect Fick Method

The two-gasometry iFM requires one arterial blood sample obtained from the radial artery and a central venous blood sample obtained from the central venous catheter collected at the same moment of TTE measurement in a heparinized syringe (0.1 mL; 1000 UI/mL), immediately processed in a blood gas analyzer. The arteriovenous oxygen difference (AVDO□) was calculated as the difference between the arterial and venous oxygen content, with the oxygen content calculated as *CaO*_2_ = (1.34 × *Hb* × *SaO*_2_) + (0.0031 × *PaO*_2_) for the arterial blood sample and *CvO*_2_ = (1.34 × *Hb* × *SvO*_2_) + (0.0031 × *PvO*_2_) for the venous blood sample, where *Hb* is the hemoglobin concentration. The CO was calculated using the formula *CO* = 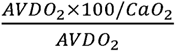.

### Statistical Analysis

Descriptive measures of categorical variables are given as the absolute frequency (n) and percentage (%). The distribution of continuous variables was assessed with histograms and Q-Q plots and the mean and standard deviation (SD) is used for normally distributed variables, and the median with interquartile range (IQR) for those with biased distributions.

The mean absolute difference (MAD) between methods is provided with 95% confidence intervals (95%CI). The correlation between CO with the Fick and TTE methods is provided as Pearson correlation coefficient and as the intraclass correlation coefficient (ICC), the latter of which accounts for repeated patient-time measurements. A linear mixed-effects model with random patient slopes was fitted to describe the change in TTE CO per unit increase in Fick CO. Bland-Altman analysis for repeated measurements^18^ was applied by fitting linear mixed-effects models with a random intercept for each patient (ID) to account for between-subject variance, a nested random intercept for each time point within a patient to account for within-subject variability across repeated observations, and a random intercept for each TTE measurement to account for intra-individual variability due to repeated TTE determinations at each time point. Models were adjusted for observer (rater performing the TTE measurements). The mean bias was obtained as the fixed effect of each model and the 95% limits of agreement (LoA) were calculated as the mean difference ± 1.96 × SD of differences, the latter derived from the combined variance components of the random effects and residual variance. All model assumptions were assessed through residual analysis and visualization of random effects from mixed-effects models through dotplots of subject-level and nested-group random intercepts.

The coefficient of variation (CV) was calculated as 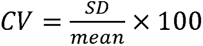 for the Fick CO and average TTE CO. The coefficient of error (CE) of TTE CO was calculated as the CV of the three CO repeated determinations per time-point divided by the squared root of 3 as described by Cecconi, et al.^19^ to obtain the precision of the reference test (TTE) as 2 times CE; 95%CI were computed through bootstrapping with the bias-corrected and accelerated (BCa) method in 10,000 re-samples. The formula described by Cecconi, et al.^19^ was used to estimate the precision of the method under study (b) by using the precision of the reference test (a):

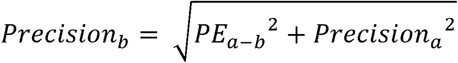

Where b is CO by Fick method, a is CO by TTE, and 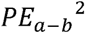 is the mean absolute percentage error (MAPE) determined with bootstrapping, and has the formula 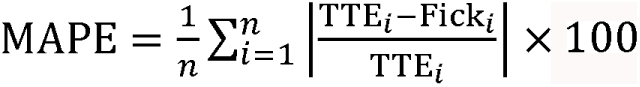. The least significant change (LSC) was calculated as 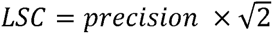 to represent the minimum change in the output measurement that corresponds to a real change in CO. All analyses and visualizations were conducted in R (v.4.5.0, RStudio v.2025.05.0). Statistical significance was defined as p = 0.05. Complete-case analyses are provided as there were no missing data.

## Results

### Descriptive characteristics of participants

A total of 243 transthoracic echocardiography cardiac output measurements were performed in 81 different time points, from a total number of 52 unique participants (**Figure 1**). The number of time points assessed per patient was 1 (n = 37), 2 (n = 8), 3 (n = 5), 4 (n = 1), and 9 (n = 1). The summary of characteristics of participants is shown in **Table 1**.

**Figure 1.**
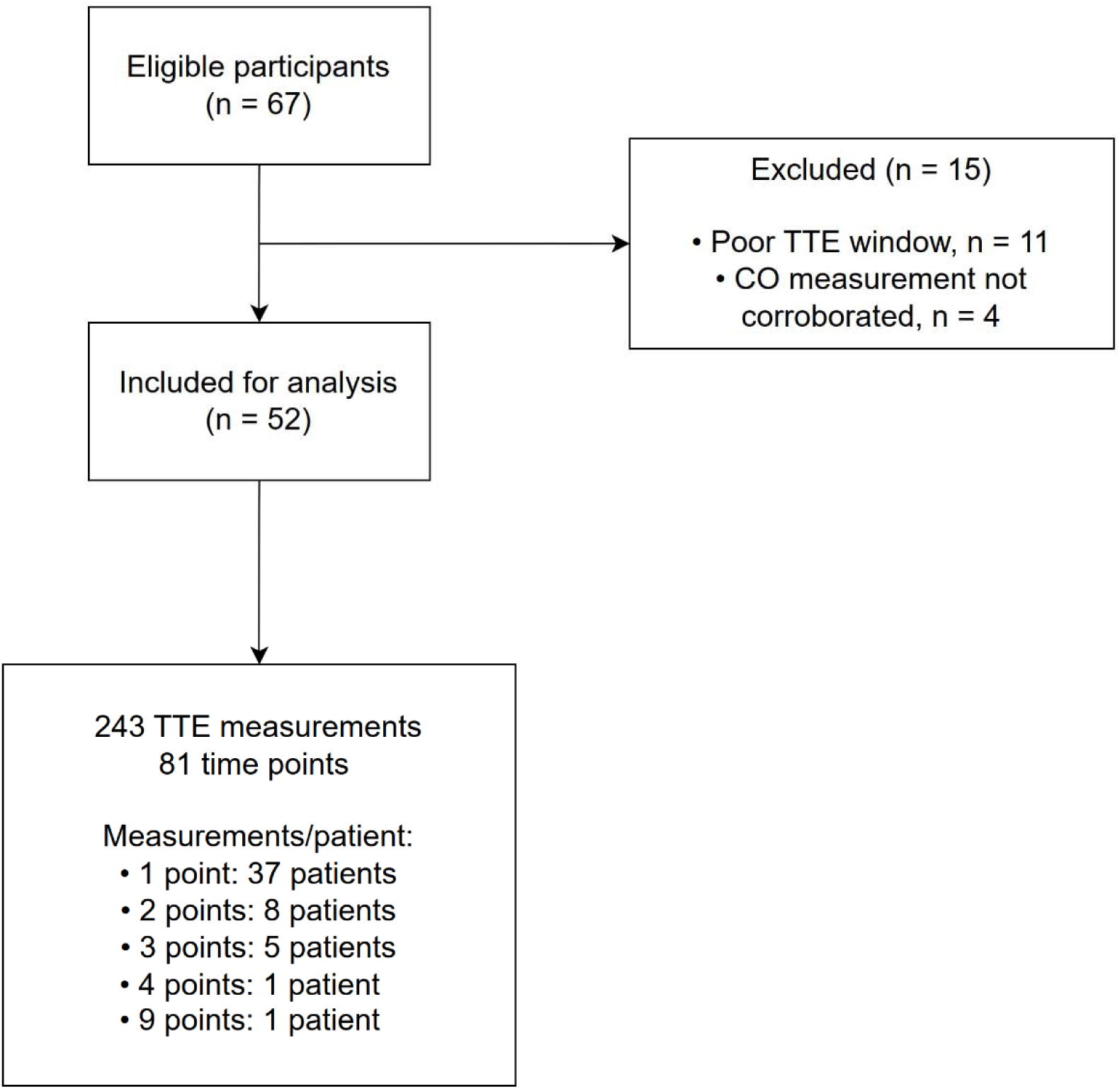
Flow diagram of participants included in the study.

### Patient status at measurement

Out of the total number of measurements, 31 (38.3%) measurements were taken while the patient was under vasopressor use. The number of measurements taken while the patient was under inotropics use was 3 (3.7%). A total of 5 and 1 participants underwent changes in vasopressor and inotropics use in between repeated measuring time points, respectively. The mean blood pressure was 83.6 (SD: 11.5) mmHg; and heart rate, 94.7 (SD: 19.7) bpm; and median FiO2, 0.33 (IQR: 0.28 - 0.4). A summary of the blood gas analysis is shown in **Table 2**.

The ventilation mode at the moment of measurement was no mechanical ventilation (n = 32, 39.5%), nonrebreather mask (n = 2, 2.5%), high-flow nasal cannula (n = 5, 6.2%), and invasive mechanical ventilation (IMV) under spontaneous (n = 5, 6.2%), CPAP|PS (n = 20, 24.7%), and ACV (n = 17, 21.0%) modes. Of the participants who were under IMV (CPAP|PS or ACV), the median PEEP value was 5 (IQR: 5 - 5.5) cmH2O; FiO2, 0.38 (IQR: 0.3 - 0.47); Pmax, 22 (IQR: 19.75 - 25); and mean Vt 377.9 (SD: 109). In those with the ACV mode, the median Pmes and DP were 22 (IQR: 16 - 23) and 14 (IQR: 12 - 17) cmH2O, respectively.

### Cardiac output with the TTE and Fick methods

The mean cardiac output with the TTE method was 6.26 L/min (95%CI: 5.73 to 6.81) and Fick method, 7.62 L/min (95%CI: 7.14 to 8.18). The correlation between the two methods was rho = 0.06 (95%CI: −0.17 to 0.27, p=0.626) and ICC = 0.05 (95%CI: −0.14 to 0.324). In the linear mixed model, there was a change in TTE CO of Fick CO of −0.004 (95% CI: −0.191 to 0.18) L/min for each unit change in mean TTE CO.

The mean absolute difference in CO between TTE and Fick was 2.76 (95%CI: 2 to 3.5) L/min. The coefficient of variation for an individual measurement of TTE was 39.5% and 31.44% for Fick. The mean CV of TTE for the repeated measurements per patient was 7.3% (95%CI: 6.2 to 8.7) and the CE was 4.2% (95%CI: 3.6 to 5), corresponding to a precision of 8.4% (95%CI: 7.2 to 10.1). The MAPE of the Fick method compared to TTE was 57.5% (95%CI: 45.5 to 74.6). The precision of the Fick method was 56.84% (95%CI: 44.34 to 74.22). The LSC was 11.9% (95%CI: 10.1 to 14.3) for TTE and 80.4% (95%CI: 62.7 to 105) for the Fick method.

The Bland-Altman plot for the repeated measures model with random effects for between-subject variance (**Figure 2A**) and within-subject variance (**Figure 2B**) shows that the mean difference (systematic bias) between TTE and Fick CO was −2.48 (95%CI: −3.82 to −1.14, p <0.001) L/min, with 95% LoA of −8.93 to 3.98 L/min in the latter model.

**Figure 2.**
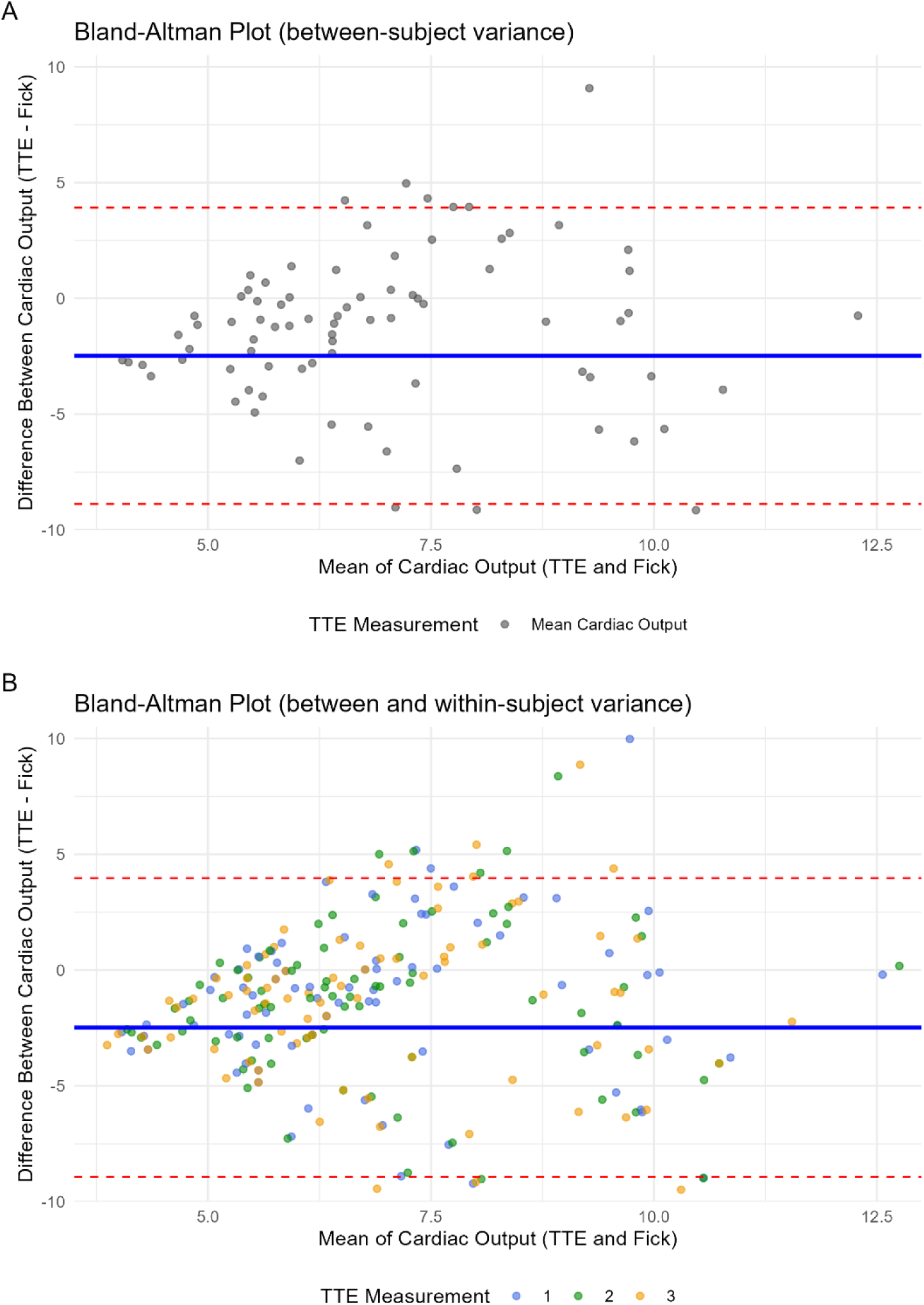
Bland-Altman plot of the difference between **cardiac output** with the transthoracic echocardiography (TTE) and Fick methods, versus the mean of both methods. Linear mixed-effects models were fitted for the differences between methods to account for the (**A**) between-subject variance due to repeated patient measurements and (**B**) within-subject variance due to repeated cardiac output measurements at the same time point. The red dotted line represents the 95% limits of agreement, whereas the blue line represents the mean systematic bias.

### Cardiac index with the TTE and Fick methods

The mean cardiac index was 3.47 L/min/m² and 4.33 L/min/m² (95% CI: 4.04 to 4.67) with the TTE and Fick methods, respectively. The correlation between the two was rho = −0.04 (95%CI: −0.22 to 0.15, p=0.637) and the ICC was −0.04 (95%CI: −0.59 to 0.26). There was a change in the Fick CI of −0.016 (95% CI: −0.247 to 0.206) for each unit increase in mean TTE CI.

The mean absolute difference of TTE and Fick CI was 1.59 (95%CI: 1.2 to 2) L/min/m². The coefficient of variation for an individual measurement of TTE and Fick CI were 35% and 33.52%, respectively, while the repeated measurements per patient of TTE had a CV of 7.3% (95%CI: 6.2 to 8.7) and CE 4.2% (95%CI: 3.6 to 5), the latter matches a precision of 8.4% (95%CI: 7.2 to 10.1). The MAPE of the Fick method against TTE was 57.5% (95%CI: 45.5 - 74.6). The precision of the Fick method was 56.84% (95%CI: 44.34 to 74.22). The LSC of TTE was 11.9% (95%CI: 10.1 to 14.3) and 80.4% (95%CI: 62.7 to 105) for the Fick method.

The mean difference (systematic bias) between TTE and Fick cardiac index was −1.53 (95%CI: −2.31 to −0.74, p < 0.001) L/min/m², with 95% LoA of −5.27 to 2.22 L/min/m² (**Figure 3**).

**Figure 3.**
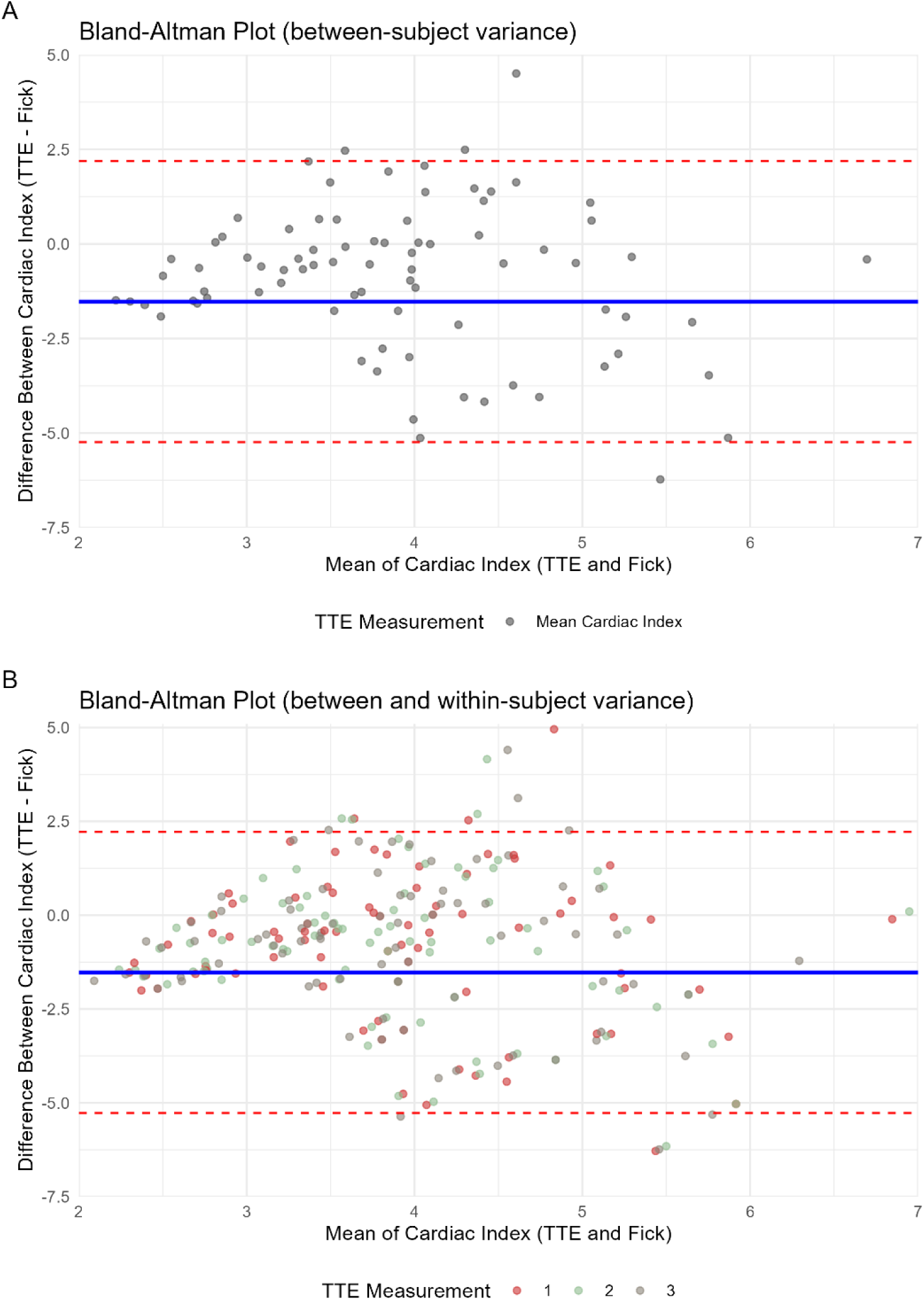
Bland-Altman plots of the difference between **cardiac index** with the transthoracic echocardiography (TTE) and Fick methods, versus the mean of both methods. Linear mixed-effects models were fitted for the differences between methods to account for the (**A**) between-subject variance due to repeated patient measurements and (**B**) within-subject variance due to repeated cardiac output measurements at the same time point. The red dotted line represents the 95% limits of agreement, whereas the blue line represents the mean systematic bias.

## Discussion

In this cross-sectional study of critically ill patients in a low-resource ICU, we assessed agreement and clinical interchangeability of CO and CI measurements from TTE (reference test) and the two-gasometry indirect Fick method (index test). We found poor agreement, with two-gasometry iFM errors far exceeding the variability expected from repeated TTE measurements. Additionally, the Fick method systematically overestimated CO and CI, risking misinterpretation of preserved cardiac output in patients who may actually require intervention.

We used three repeated TTE measurements to determine reference test precision, following the precision-versus-measurements curve by Cecconi et al.^19^ This number of measurements was considered to yield acceptable precision for our study. In the past, a fixed precision of 30% has been used for cardiac output studies using intermittent dilution as the reference test.^24^ Nonetheless, determining precision in the context of the study population for validation is currently preferred.^19^ In our study, TTE precision was 8.4% (95%CI: 7.2–10.1) and the LSC was 11.9% (95%CI: 10.1–14.3), meaning changes below this threshold may reflect measurement error. Said differently, a change in CO or CI of 11.9% or more is needed to be confident that there is a real change in patient status. This aligns with the 11% LSC reported by Jozwiak et al. for three repeated TTEs in patients with sinus rhythm.^25^ In contrast, the two-gasometry iFM had poor precision (56.8%, 95%CI: 44.34 to 74.22) and LSC of 80.4% (95%CI: 62.7 to 105), implying that any change in CO or CI below 80.4% is not clinically meaningful and likely due to measurement error. The elevated MAPE (57.5%, 95%CI: 45.5–74.6) further highlights the substantial inaccuracy of the Fick method compared to TTE.

We included repeated measurements at different time points for a subset of patients to model nested intra-individual variation due to changes in clinical status. Most variation was explained by inter-individual differences, followed by within-individual changes, while variation from repeated TTEs at the same time point was minimal. All models were adjusted for observer. This design enabled robust estimation of systematic bias and limits of agreement. For CO, the mean bias was −2.48 L/min (95% LoA: −8.93 to 3.98), and for CI, −1.53 L/min/m² (95% LoA: −5.27 to 2.22), both with wide limits. Practically, this indicates that the Fick method overestimates CO and CI by ∼2.5 L/min and 1.5 L/min/m², which could result in undertreatment of patients with reduced cardiac function. Since TTE itself underestimates CO by ∼0.64 L/min versus thermodilution,^17^ the true bias of the Fick method may be lower. Still, the wide limits of agreement confirm that the Fick method is not interchangeable with TTE for CO and CI assessment.

We observed poor correlation between the two methods. The linear mixed-effects model showed no predictable change in TTE CO per unit increase in Fick CO. In these regressions, we used the average of the three TTE measurements as the predictor and the Fick method as the outcome to avoid regression dilution,^26^ which occurs when error exists in the independent variable, contrary to error in the dependent variable which causes unbiased estimates with wider intervals.^27^ A beta of 1 would indicate equivalence (1 L/min change in TTE CO equals 1 L/min change in Fick CO), allowing for bias correction. However, we found near-zero coefficients (–0.004 L/min for CO; –0.016 L/min/m² for CI) and poor correlation, indicating that correction for systematic bias is not appropriate due to lack of predictive value.

Similarly, Sánchez Velázquez evaluated CO estimates from two AVDO□-based formulas against thermodilution in 54 ICU patients (540 measurements). Correlation coefficients were low (0.15 and 0.33), and mean biases were 0.001 L/min and –1.4 L/min, respectively, further supporting that such formulas are not reliable substitutes for thermodilution in critically ill patients.^10^

De la Cruz et al. compared CO measurements obtained via thermodilution, transesophageal bioimpedance, and the previously described equation (CO = 125 × BSA/8.5 × AVDO□). Notably, VO□ was neither measured directly nor estimated indirectly, but assumed by the authors without accounting for patient-specific conditions. Mean CO values were 10.2 + 3.8 L/min (thermodilution), 10.4 ± 3.6 L/min (bioimpedance), and 6.2 ± 1.3 L/min (two-gasometry equation), with the latter significantly lower (p < 0.05). Although no correlation analysis was conducted, the substantial discrepancy between the two-gasometry method and both reference techniques highlights its inaccuracy.^28^

In addition to its lack of correlation with CO, the two-gasometry method has several drawbacks. It requires frequent arterial punctures, which can cause pain, thrombosis, aneurysms, nerve injury, and arteriovenous fistulas,^29^ or necessitate arterial line placement for repeated sampling—raising both costs and the risk of complications. A systematic review estimated bloodstream infection rates from arterial lines at 1.7 per 1000 device-days, comparable to non-tunneled central venous catheters (2.7) and higher than peripheral venous catheters (0.5, 95%CI: 0.2–0.7).^30^ Reported vascular complication rates for radial artery catheters include 19.7% overall, with 0.1% temporary ischemia, 14.4% permanent ischemia, 0.5% hematoma, and 0.1% pseudoaneurysm.^31,32^ Each blood gas analysis costs approximately 37.25 AUD (∼23 USD). At least two samples are ordered to estimate CO with the iFM and technical failures may lead to performing more than one arterial puncture.^33^ Lastly, central venous catheter use and line manipulation for sampling carry a risk of bloodstream infection.^30^

Strengths of this study include the use of TTE as the reference test, a widely validated and recognized method for CO measurement in critically ill patients. The statistical analysis was robust, accounting for repeated measures and both intra- and inter-individual variability. Being a single-center study avoided inter-observer variability across institutions.

The main limitation was the absence of a gold standard, such as thermodilution via PAC, which would have enabled assessment of the true accuracy of the two-gasometry iFM. However, PAC is invasive, carries risks, and is often unavailable in resource-limited settings. Another limitation was the lack of direct VO□ measurement, a key component of the Fick principle, though assumed VO□ values are common in clinical practice. The sample size was modest, and although we did not assess inter-observer agreement, we adjusted for observer effects in our analysis.

These findings are especially relevant to resource-limited hospitals, where access to complex methods like PAC thermodilution may be lacking. However, the high variability of the two-gasometry iFM emphasizes the need to prioritize TTE as a more reliable and reproducible alternative for hemodynamic assessment. Still, caution is warranted when applying these results to other settings, as they assume the availability of trained sonographers and echocardiography equipment, both of which may be limited in such environments.

## Conclusion

Cardiac output measured by the two-gasometry indirect Fick method shows poor agreement and clinical interchangeability with TTE. Due to its low precision, a change of at least 80.4% in CO or CI is required to reflect a true change in patient status. The two-gasometry iFM systematically overestimates CO and CI by ∼2.5 L/min and 1.5 L/min/m², potentially leading to undertreatment in patients with impaired cardiac function. Given its high bias, poor precision, and wide limits of agreement, the two-gasometry iFM is not suitable for clinical decision-making in critically ill patients.

## Ethics Statement

Procedures were followed in accordance with the ethical standards of the institutional responsible committee and with the Helsinki Declaration of 1975. Written informed consent was obtained from all participants. This study was approved by the ethics and research committee of the Health Services of the State of Queretaro, Mexico on 14/02/2024 under study title “Método de Fick vs ecocardiografia funcional como método de monitoreo del gasto cardiaco en pacientes de la unidad de cuidados intensivos del Hospital General San Juan del Rio” (registration number: 1616/08/09/2024).

## Data Availability

The data that support the findings of this study are openly available in the Harvard Dataverse at https://doi.org/10.7910/DVN/J4ONSU.

## Code Availability

The code documenting the analysis in this study is openly available in https://github.com/javimangal/cardiac-output-fick

## Acknowledgements

None.

